# Multimodal CNN Models Allow for the Accurate Classification and Grading of Preoperative Meningioma Tumors

**DOI:** 10.1101/2023.03.15.23287326

**Authors:** Mihir Rane

## Abstract

Magnetic resonance imaging (MRI) and computed tomography (CT) scans are vital for diagnosing brain tumors, but human error, image subtleties, cyst growth, and nuances in World Health Organization (WHO) grading can impede accuracy. Invasive biopsies remain the only definitive method for meningioma diagnosis. Convolutional Neural Networks (CNNs), machine learning models used in image classification, offer a promising solution. By fine-tuning the pre-trained CNN EfficientNetB0 on various preoperative brain tumors and meningioma subtypes, image-based diagnosis can become more robust and accurate. In this study, two CNN models either classified or graded multimodal CT and MRI images. One dataset included tumor types (meningioma, glioma, pituitary, cysts, or none), while the other had images WHO graded one to three. The data, from accurately annotated and diverse open-source databases, was normalized, augmented, and stripped of excess information. Additionally, class-average and Focal Tversky Loss were included to assess and reduce incorrect outputs. Results were analyzed using accuracy, f1, recall, precision, loss, confusion matrices, Receiver Operating Characteristic (ROC) analysis, and attention studies. Both CNNs achieved over 98% accuracy with high recall and precision scores. ROC area under the curve (AUC) scores above 0.978 indicated strong class discrimination. The attention study indicated focus on tumor mass instead of extraneous variables. Multimodal CNNs, particularly the EfficientNetB0 model, are potential alternatives to invasive biopsies and human evaluation. Their capability to handle complex meningioma cases suggests promising avenues for other tumor types or diagnostic modalities at a cheap cost.

## 1. INTRODUCTION

Meningiomas are the second most common type of primary brain tumor, accounting for around thirty percent of all central nervous system (CNS) tumors, and impacting around one in one thousand individuals worldwide^12,28,34^. Typically noncancerous in early stages, meningiomas occur in the meninges, the membranes surrounding the brain and spinal cord^7,28,34^. Monosomy 22 and inactivating mutations of NF2 have been linked to meningioma, although the pathophysiology and etiology are not well-known^7,33^. Depending on the tumor’s location, meningioma can cause seizures, loss of sensory function, memory loss, motor loss, and other neurodegenerative issues^7,28^. Therefore, despite being typically slow-growing and non-cancerous, physicians recommend immediate surgery and resection before malignancy or the rare case of metastasis^10^.

Diagnostic testing for meningioma growth and grading include magnetic resonance imaging (MRI), computed tomography (CT), cerebral angiograms, and biopsy. Often, multiple diagnostic tests are necessary for an accurate diagnosis^15^. Structural MRI and CT scans have become the most common diagnostic methods for brain tumors, providing clear visual images for examining patient brain structure^11,33^. Additionally, MRI scans themselves have multiple modalities, most commonly T1-weighted, T2-weighted, and Fluid Attenuated Inversion Recovery (FLAIR). These modalities manipulate the repetition (TR) and echo (TE) time variables, each aiming to increase the highlighting of neoplasms–abnormal growths^2,8,30^. Furthermore, contrast agents like C+ and Gadolinium and different cross sections (Sagittal, Coronal, or Axial) are employed in both MRI and CT scans in order to highlight vascular abnormalities, tumors, and inflammation^9,29^.

After classification, meningiomas are assigned a specific World Health Organization (WHO) grade, depicting the appearance and abnormalities of cancer cells^12,18,23^. Grading is typically done through histopathological means under a microscope after biopsy, based on mitosis rates, specific features, and metastasis^18^. However, grading can also sometimes be accomplished through extensive image evaluation^11,16^. Incorrect grading of meningioma remains a large issue due to overlapping features and inherent subjectivity in visual assessment. Assigning the wrong grade can be detrimental, as it may result in inappropriate treatment plans^13,16,23^. For this reason, although biopsies carry risks such as subdural hematomas, infections, strokes, and blood clots, practitioners still rely on this medium for conclusive diagnoses^14^. Fortunately, recent advancements in machine learning and deep learning presents a promising avenue to mitigate human subjectivity, reduce biopsy risks, and streamline the diagnostic process^5,22^.

Image classification is most accurately and commonly performed using Convolutional Neural Networks (CNNs)^4,27^. However, current CNN models struggle to accurately classify and grade meningiomas compared to other brain tumors, a phenomenon that can be attributed to the immense diversity within meningioma features. There is not enough training data for easy generalization across scenarios^13,16,23^. This highlights the need for larger and more diverse datasets, specifically for meningiomas, in order to improve CNN performance in this area. Employing multimodal CT and MRI scans with various cross-sections would provide a richer dataset for training CNN models. The enhanced information from different modalities enables better differentiation of subtle abnormalities and variations in tissue characteristics. With the use of multimodal data, a Convolutional Neural Network will be able to accurately differentiate meningioma from other preoperative brain tumors, while separately grading the meningioma tumors. Such an advent would reduce the need for invasive biopsies while maintaining a cheaper and quicker form of diagnosis.

Due to the employment of CNNs, the study will not include non-imaging data such as patient medical histories, genetic information, or blood tests in the CNN model. Other machine learning approaches outside of CNNs, such as support vector machines or random forests, will not be explored in detail. Additionally, the study will not track patient outcomes over time to assess the long-term efficacy of the CNN model in clinical practice. Furthermore, while multimodal data will be augmented, limited training data pertaining to meningioma will affect the model’s training and generalization capabilities. Imaging datasets may also contain biases related to patient demographics, scanner types, and imaging protocols, leading to skewed results and affecting the model’s applicability across different patient populations.

This research is grounded in the theoretical framework of machine learning models, particularly leveraging Convolutional Neural Networks (CNNs) for the classification and diagnosis of meningiomas. CNNs are Feed-Forward Neural Networks that process inputs through non-linear functions like Sigmoid or ReLU, with weights adjusted via gradient descent and bias values optimized^3,17,31^. The convolution layer, the core of CNNs, performs dot products between a kernel and the receptive field, creating activation maps^6,26^. Pooling layers reduce spatial size and computational load, and fully connected (FC) layers illustrate input-output relationships^22,25,26^.

The study utilizes EfficientNetB0 as the backbone of the CNN, chosen for its compound scaling approach, which balances depth, width, and resolution. EfficientNetB0, with its pre-trained weights from the ImageNet database and over 11 million trainable parameters, handles large-scale image data effectively. Its architecture includes a GlobalAveragePooling2D layer, a dropout layer to mitigate overfitting, and a final dense layer using the softmax function for classification. Attention mechanisms in EfficientNetB0 enhance the model’s ability to process images accurately^20,31^. Callback functions like TensorBoard, ModelCheckpoint, and ReduceLROnPlateau monitor and optimize the training process, further preventing overfitting and improving model performance^1,20^. This framework provides a robust methodology for automating meningioma detection and grading.

Regarding methodology, the model utilized a dataset combining previously documented data and new cases from various global hospitals, involving MRI and CT images. To address overfitting and enhance the multimodal dataset, data augmentation techniques such as mirroring, scaling, rotations, and contrast adjustments were applied. Image preprocessing included steps like image registration, bias field correction, skull stripping, and normalization. Training strategies involved using class-average loss and Focal Tversky loss with accumulated gradients for effective batch processing. The model’s performance was evaluated using common metrics such as f1 score, recall, precision, accuracy, confusion matrices, Receiver Operating Curve (ROC) analysis, and a model attention study to discover if the models are truly focused on the tumor mass itself.

## 2. RESULTS

### 2.1 Implementation Details

Results were obtained using an HP desktop: Intel®Core™i9-9900K CPU @ 3.60 GHz, 16.0 GB of RAM @2666 MHz, 512 SSD, 2 TB HDD, and NVIDIA GeForce RTX 2080 Ti GPU. Implementation was done in Python using TensorFlow v1.13.1 with Keras, and PyTorch lightning v0.7.3 with PyTorch back-end v1.3 on JupyterNotebook via Visual Studio Code.

### 2.2 Overall Performance Study

**Table 1:** Performance summary for each tumor type averaged across five folds.

| Tumor Type | F1 | Recall | Precision | Accuracy |
| --- | --- | --- | --- | --- |
| No Tumor | 97.55 ± 1.36 | 96.89 ± 1.51 | 98.21 ± 1.23 | 99.21 ± 0.54 |
| Meningioma Tumor | 97.16 ± 1.29 | 95.59 ± 1.12 | 98.78 ± 1.48 | 99.60 ± 0.23 |
| Pituitary Tumor | 96.76 ± 1.20 | 95.54 ± 1.07 | 98.01 ± 1.33 | 98.82 ± 0.66 |
| Glioma Tumor | 97.74 ± 1.24 | 97.23 ± 1.24 | 98.25 ± 1.55 | 99.31 ± 0.44 |

**Table 2:** Performance summary for each meningioma grade averaged across five folds.

| WHO Meningioma Grade | F1 | Recall | Precision | Accuracy |
| --- | --- | --- | --- | --- |
| Grade I | 97.76 ± 1.08 | 96.83 ± 1.03 | 98.71 ± 1.14 | 99.19 ± 0.57 |
| Grade II | 98.17 ± 1.17 | 97.71 ± 1.33 | 98.64 ± 1.01 | 99.24 ± 0.34 |
| Grade III | 97.78 ± 1.10 | 97.24 ± 1.00 | 98.33 ± 1.20 | 99.06 ± 0.43 |

The overall performance study indicates high quality and reliability of both classification tasks in this work. The accuracy reached extremely high scores, specifically >98.82% for all classes. The high precision and recall values furthermore indicate high quality of the model. Average precision scores ranged from 98.01% to 98.78% in the tumor classification task and 98.33% to 98.71% in the sub-grading task. These results indicate very few false positive values and the return of more relevant results than negative. Average recall scores range between 95.54% to 97.23% in the classification model and 96.83% to 97.71% in the sub-grading model, indicating the rare return of a false negative value. High recall scores are essential to studies in a medical context due to the immense risk involved in mistaking a patient with a tumor as healthy.

Regarding the pituitary tumor subtype in the classification task, the diffuse nature of the tumors and the less defined gradients are probable explanations for the lower classification performance. For the meningioma category, the reason for the lower recall values can be attributed to the larger number of small tumors (<3ml) compared to other subtypes. In addition, outliers have been identified in this dataset where a small amount of the tumors were enhanced due to calcification.

While tumor volumes and outlier MR and CT scans are reasons for the discrepancy in values across the board, the nature of the convolutional neural network architecture and training strategy used can further explain those results. Given GPU memory limitation, the preprocessed MR and CT scans have undergone a significant down sampling, and as such small tumors are reduced to very few voxels, impacting performance across the board.

### 2.3 Accuracy and Loss Analysis

The data analysis reveals that high accuracy and AUC scores were achieved during an upwards trend through 12 epochs during the validation and training steps of the model. Below are tables listing the Area Under the Curve (AUC) scores of the classification and grading tasks demonstrated in figure 4.

**Figure 1.**
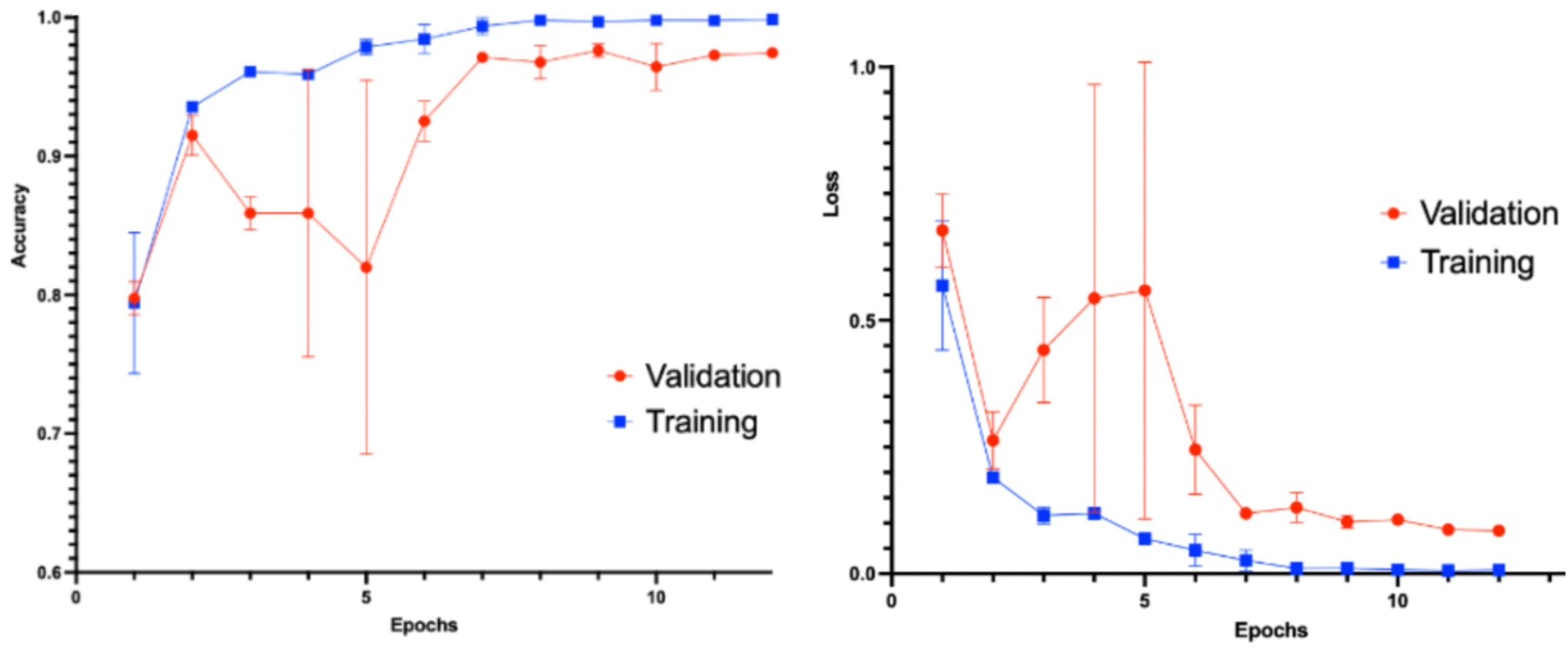
Graphs of the validation and training accuracy (left) and loss (right) averaged over five splits during 12 epochs for the classification model. Error bars represent a 95% confidence interval, while the solid line represents the average.

**Figure 2.**
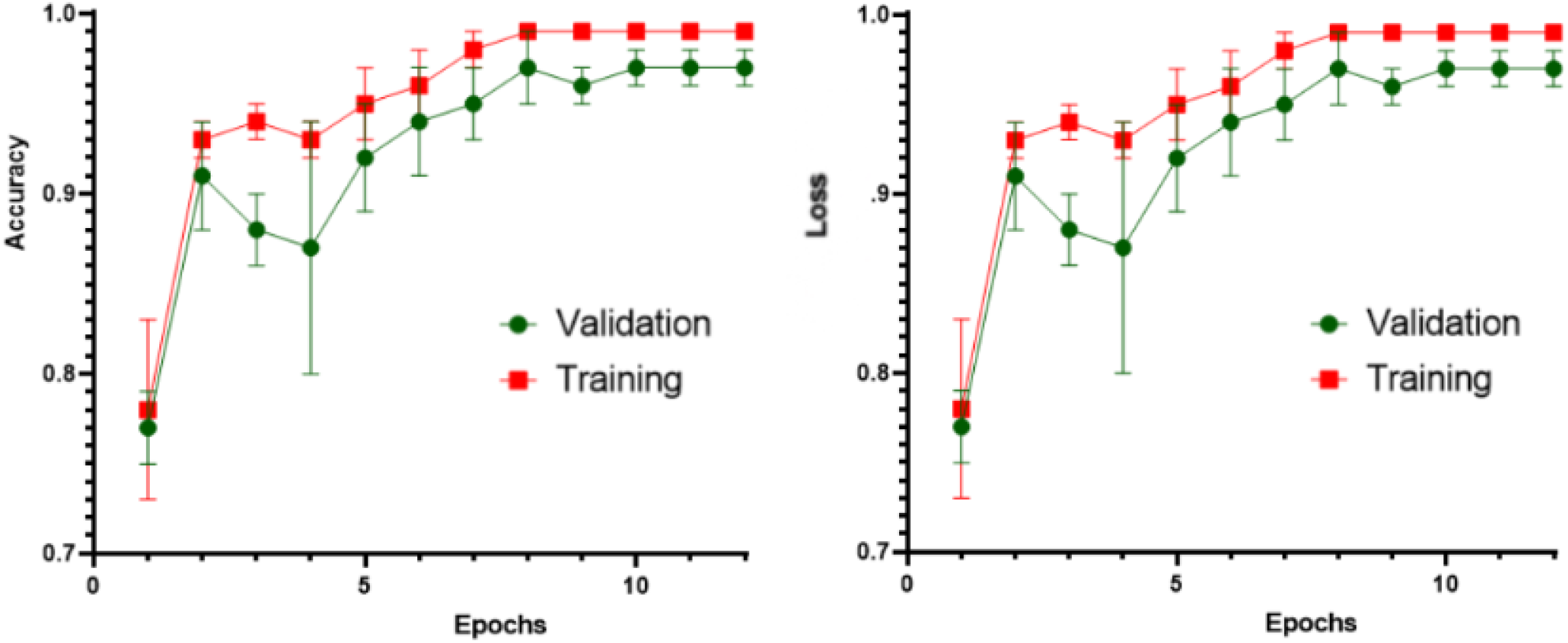
Graphs of the validation and training accuracy (left) and loss (right) averaged over five splits during 12 epochs for the grading model. Error bars represent a 95% confidence interval, while the solid line represents the average.

**Figure 3.**
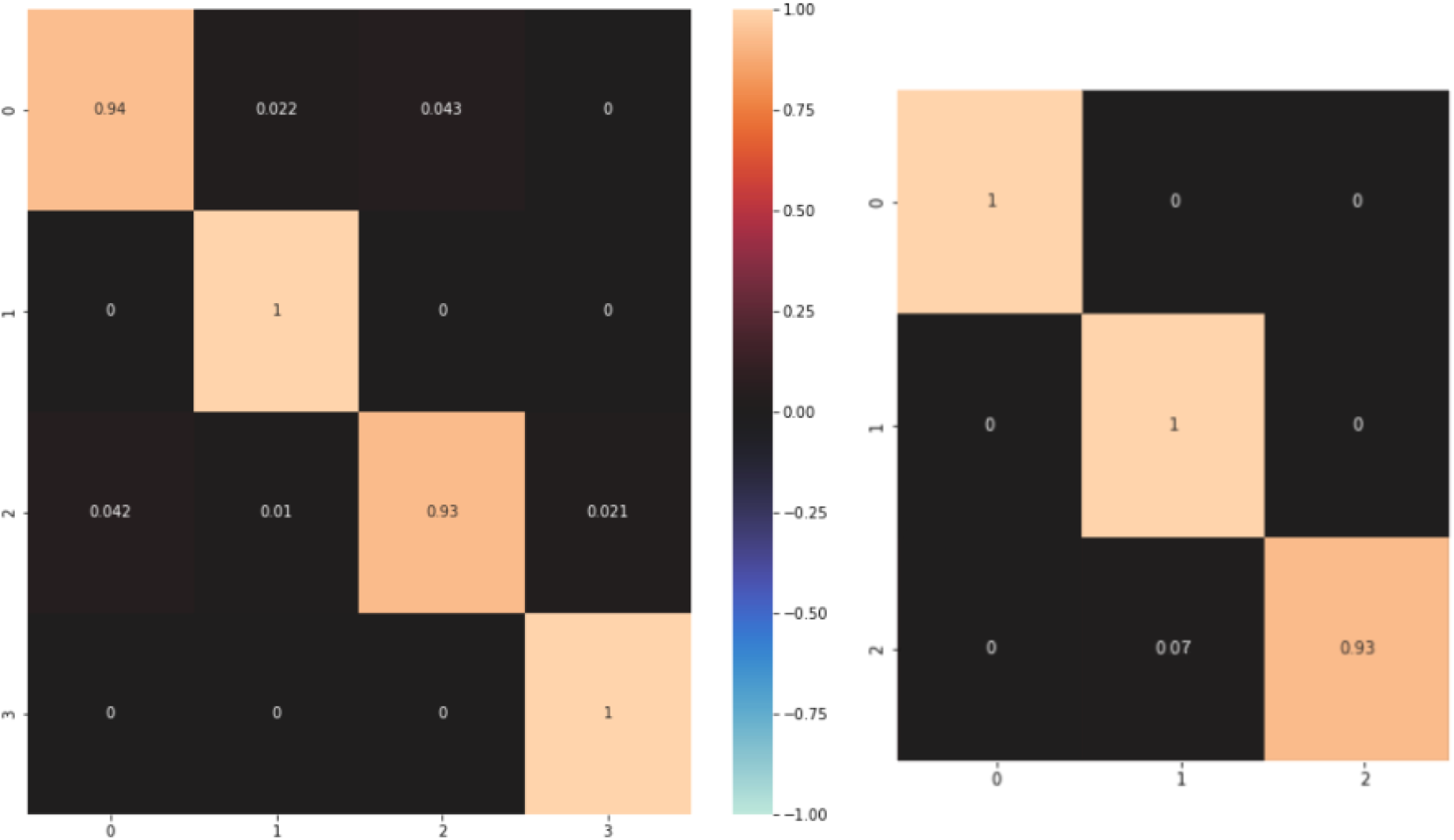
Heat maps of the confusion matrix regarding accuracies of tumor classification (left) and meningioma tumor grading (right), with 1.00 representing the highest accuracy.

**Figure 4.**
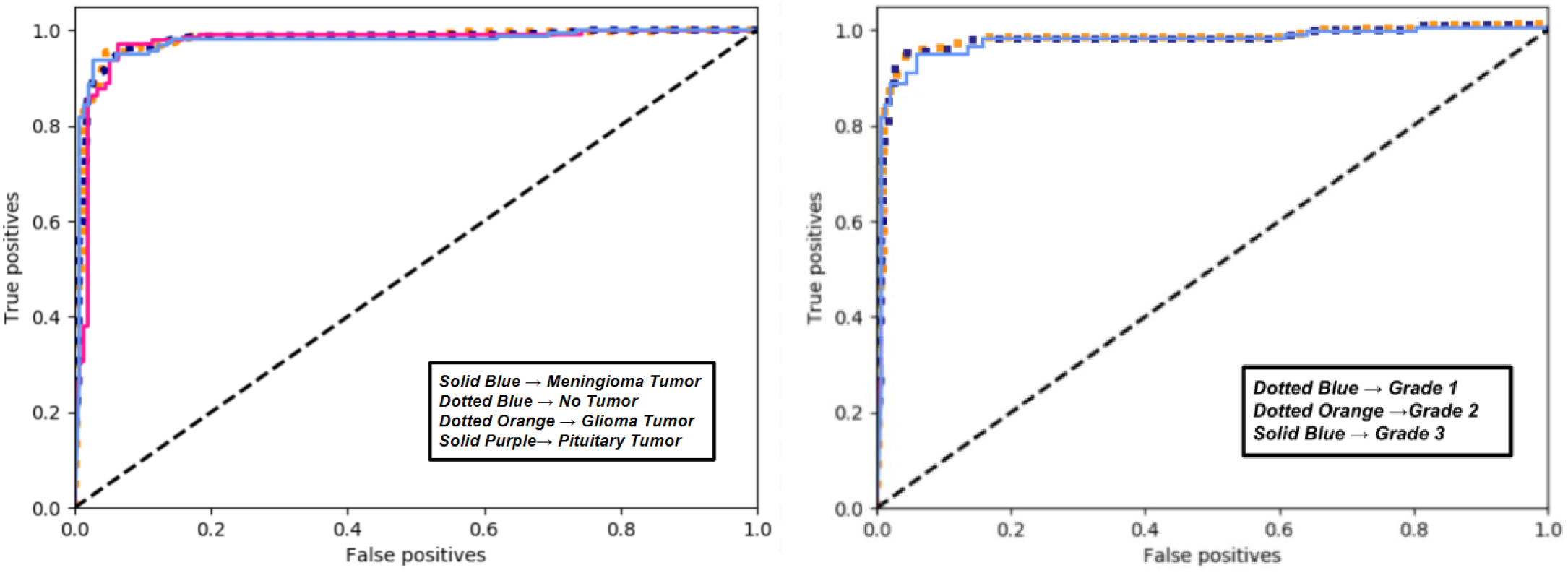
Receiver Operating Characteristic (ROC) curves depicting false positive versus true positives rates given threshold values between 0 and 1 with intervals of 0.2 for the classification task (left) and grading task (right). The graphs were generated using the SKlearn utility in python. A baseline of 0.5 Area Under the Curve (AUC) is demonstrated by the black line.

**Figure 5.**
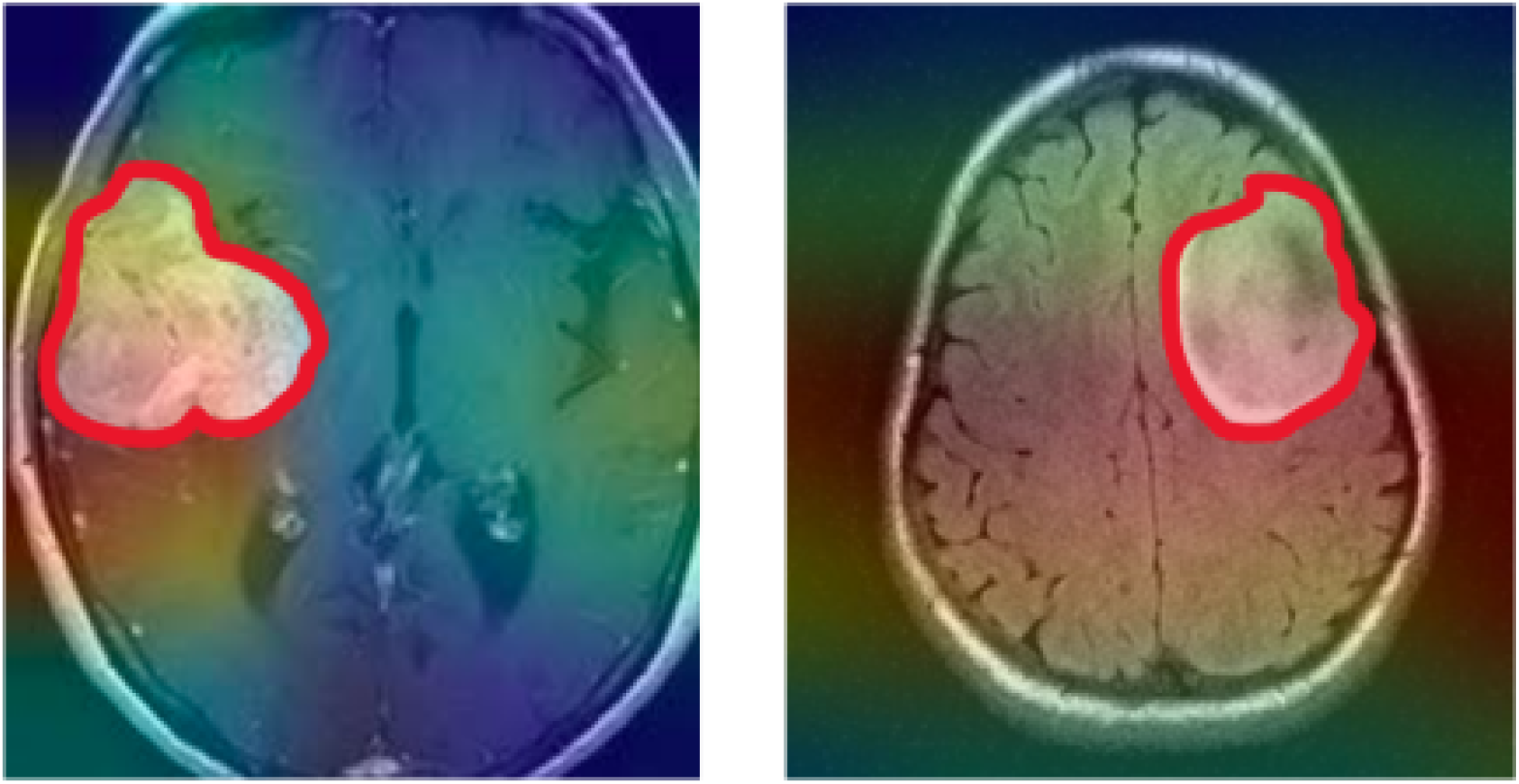
Images taken with GradCam TensorFlow extension, showing a heat map of the areas with the highest attention in two T1-weighted MRI images. Highest attention approaches red from blue up the color scale. Manual delineation is shown in solid red. These samples are unprocessed and are used only as examples post-training. Therefore, the images are not skull-stripped.

Classification Task

|  | Meningioma Tumor | Pituitary Tumor | Glioma Tumor | No Tumor |
| --- | --- | --- | --- | --- |
| AUC Score | .987 | .981 | .982 | .985 |

Grading Task

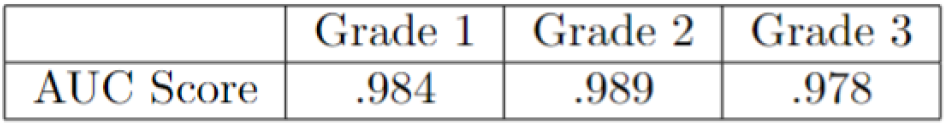

Grading Task:
|  | Grade 1 | Grade 2 | Grade 3 |
| --- | --- | --- | --- |
| AUC Score | .984 | .989 | .978 |

In concordance to the high accuracies revealed in previous tests, both models demonstrate exemplary AUC scores. During classification, the models were able to accurately distinguish positive and negative classes. Furthermore, AUC was calculated using a Receiver Operating Characteristic (ROC) in order to include variables such as threshold-invariance and scale-invariance, two variables absent if accuracy is used alone. The positive instance is ranked higher than the negative instance on both CNNs more than 98% of the time, indicating high quality of the model, the quality of the loss functions, and its reliability to a practitioner.

The confusion matrices corroborate the results shown in the ROC curve. True-positive classes reach high accuracies, ranging from 93% up to 100% in the classification task and grading model. In both models, the extremes are 0, furthermore indicating high quality and stability. The Confusion Matrix displays the tendency to rank positive instances higher than negative instances almost all the time.

As described earlier, some discrepancies and false-positives seen in both models can be attributed to tumor mass less than 3 ml. The slightly depressed percentage of true-positives could also be due to neoplastic growth included in the dataset that are not tumors, like cysts. Additionally, the validation accuracies range greatly and are slightly lower than the training accuracies for each epoch. There are two plausible explanations for this discrepancy: slight overfitting of the model or data splits drastically affecting validation performance. The latter instance proves more likely due to the standard deviation (SD) of the validation step ranging greatly. Overfitting is less likely due to the high quality and stability of the training step.

### 2.4 Attention Study

Attention, allocated importance during classification, in the images seem to surround the manually marked tumor mass, with the model giving most importance to these contrasting sections. This study rules out any underlying possibility of pseudo-accuracy generated from falsified attention to extraneous variables, such as barcodes or markings outside the brain mass itself. Attention patterns in this study corroborate the high accuracies noted previously.

Although attention is fairly concentrated in the image on the left, attention seems to spread across the 12 middle of the MRI image on the right. This discrepancy can be attributed to displacement of neighboring brain mass in response to the growth of a foreign mass. Another explanation for superfluous attention would be the inclusion of outlier non-cancerous cysts to the training and testing datasets. Cyst growth has been attributed to excess brain deformation when compared to tumor growth. Small amounts of attention could have formed due to patterns in tumor localization as well. Another explanation for the discrepancies could be due to the use of unprocessed data for the sake of the attention study.

## 3. DISCUSSION

This study trained convolutional neural networks (CNNs) from various CT and MRI modalities to classify and grade meningioma tumors in the brain. The dataset comprised images from multiple scanners across hospitals worldwide. Using an EfficientNetB0 backbone, along with data augmentation and preprocessing, the classification and sub-grading models for pituitary tumors and meningiomas demonstrated high quality and performance, with average precision scores ranging from 98.01% to 98.78% and average recall scores between 95.54% to 97.23%. The high recall scores demonstrate low return of false negatives, which are detrimental in a medical context. Moreover, despite the challenges posed by small tumor volumes and outliers, the models achieved accuracy and AUC scores above approximately 0.98, indicating a reliable distinction between positive and negative classes. Furthermore, the attention study with GradCam confirms the model’s focus on relevant tumor areas, supporting the high accuracy and stability observed.

The findings highlight the potential of CNNs to enhance neurosurgical and radiological practices by accurately diagnosing and grading brain tumors. By analyzing CT and MRI images with varying contrasts, temporal sequences, and positions, CNNs can improve in diagnostic accuracy and assist in surgical planning. Despite introducing variability, the high accuracy of CNN models in this study displays the efficacy of multimodal data in improving datasets. The accuracy of the lightwork EfficeientNetB0 model also exemplifies the feasibility of integrating machine learning into medical practices globally, including in resource-limited settings.

This study aimed to evaluate the effectiveness of CNNs in medical image analysis and demonstrate their potential in clinical applications. It met its objectives by achieving high overall scores, demonstrating high discrimination capabilities, and allocating attention to areas of importance. The use of a diverse and extensive multimodal dataset and various image augmentation and preprocessing techniques further supported the goal of creating a robust and reliable model for diagnosing and grading meningioma tumors.

Future work should focus on refining CNNs with false-positive, true-negative, and more complex meningioma samples to enhance practical classification and identify specific images that lead to erroneous predictions. Expanding the dataset to include various other types of neoplasms would increase robustness. Additionally, incorporating modalities like histology and angiography in separate machine learning models could improve accuracy. Experimentation with different environments, including those with more limited GPU resources, is necessary to evaluate the model’s applicability in low-resource settings. Modifying callback and loss function parameters and adjusting convolutional and pooling layers in an ablation study would help address issues with miniature tumor growth or indentations smaller than 3 ml. Additionally, distinct models could be fine-tuned to assess the growth grades of other tumor classifications, such as glioma and pituitary, which were included in this study.

The study faced challenges related to the high memory footprint of high-resolution MR and CT images, even when using advanced GPU environments. Down-sampling was necessary, which may have impacted some aspects of the model’s performance. Additionally, neoplasms less than three milliliters and outliers such as cyst growth may have caused confusion within the model. However, through these challenges, both models performed exceptionally.

CNNs have a promising potential in the medical field, particularly for diagnosing and grading brain tumors. The success of the EfficientNetB0 architecture in maintaining high accuracy, precision, and recall scores illustrates the feasibility of integrating machine learning into clinical practice. Future research and experimentation will be essential to refine these models, expand their applications, and ensure they are accessible and effective in diverse healthcare settings globally. This study is a foundational step towards more sophisticated and impactful machine learning in medicine and radiology, allowing for excellent patient safety and accurate diagnoses.

## 4. MATERIALS AND METHODS

The current observational study records the efficacy of both CNN models using a variety of common scoring and evaluation metrics that will be discussed later. Validation loss in specific was observed during the training step. However, it is primarily important to understand the sample employed. The study itself is cross-sectional, as the population consists of a variety of individuals with various tumor classifications. Additionally, the image data itself is based on a multimodal approach to diagnosis.

### 4.1 Dataset

The dataset used in this research was an amalgamation of previously documented datasets as well as data from a variety of documented cases from both inpatients and outpatients in public hospitals and private practitioners. The MRI images were taken from both 1.5T and 3.0T scanners and the CT images were taken by special X-ray scanners at 12 different hospitals and institutions in the United States, Europe, and India. All data from CT and MRI are grayscale and with only one channel. Patient age ranges from 8 years to 82 years, gender includes 54.8% female and 45.2% male, and racial demographics are unknown. Training and testing splits for the tumor classes were conducted randomly from 7-17% in order to make up for the unbalanced data regarding “No Tumor” and to avoid overfitting. Random splits carried on for the grading model in order to replicate a similar situation to the classification task. From there, the validation split was 20% of the training data. Due to the constant splits, total validation samples are not shown.

**Table 1:** A table illustrating the number of sample images used for each type of tumor in the tumor classification model before data augmentation. Images are from multiple modalities. Training and testing data amounts are reported.

|  | Meningioma | Glioma | Pituitary | No Tumor |
| --- | --- | --- | --- | --- |
| Training Data | 822 | 826 | 827 | 395 |
| Testing Data | 115 | 100 | 74 | 105 |

**Table 2:** A table illustrating the number of sample images used for each WHO grade of meningioma in the sub-grading model before data augmentation. Images are from multiple modalities. Training and testing data amounts are reported.

|  | Grade 1 | Grade 2 | Grade 3 |
| --- | --- | --- | --- |
| Training Data | 1286 | 1278 | 1282 |
| Testing Data | 201 | 202 | 197 |

**Figure 6.**
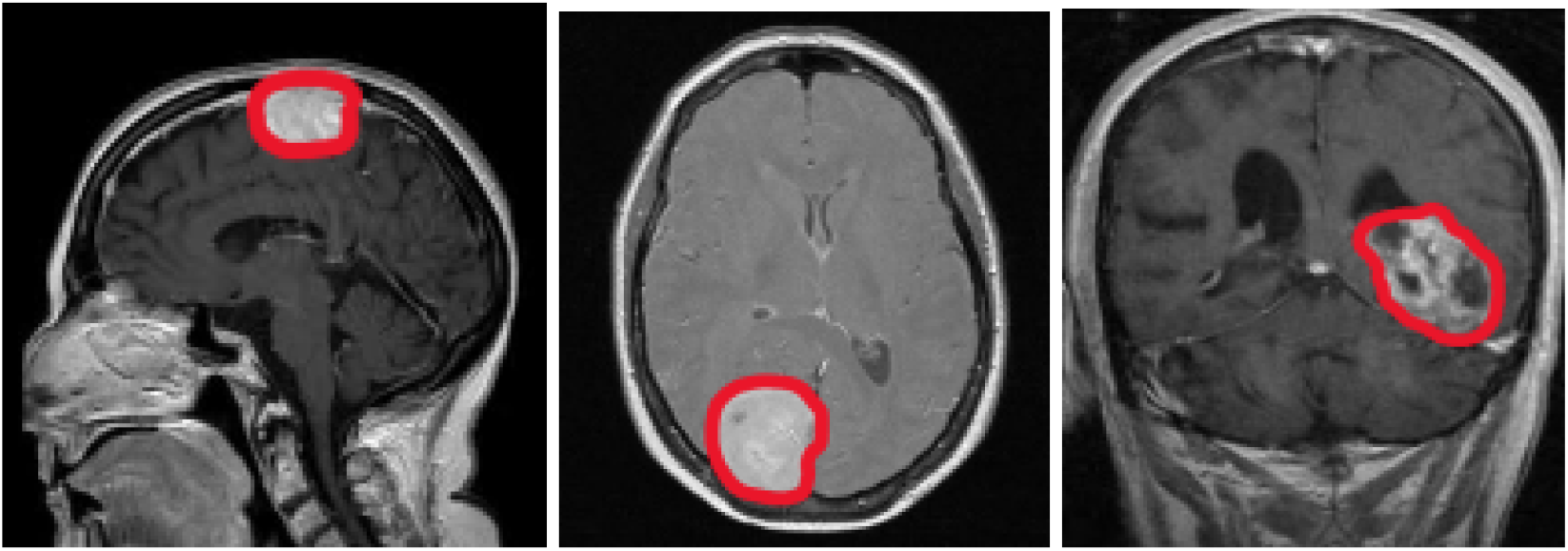
Examples of brain tumors from the raw MRI volumes collected in this study. All images are representations of meningioma tumor growth. The far left is an axial cross-section, the middle is a sagittal section, and the far right is a coronal section of meningioma. Images were not altered. For the purposes of this figure, the tumor masses were manually annotated in red. Annotated images were not used in the training steps of this model.

All data collected are preoperative, before treatment such as surgery, chemotherapy, radiotherapy, or other therapeutic measures. Overall volume of tumor resection was greater than 1ml for those surgical patients. MRI and CT volume dimensions cover [192; 576] × [240; 640] × [16; 400] voxels, and the voxel size range [0.34; 1.17] × [0.34; 1.17] × [0.50; 8.0] mm^3^. These values include all tumors in the classification model’s datasets. In the meningioma grading dataset, MRI and CT volume dimensions cover [192; 512] × [224; 512] × [11; 290] voxels, and the voxel size range [0.41; 1.05] × [0.41; 1.05] × [0.60; 7.00] mm^3^. For reference, an average MRI or CT volume is [349 × 363 × 85] pixels with a spacing of [0.72 × 0.72 × 4.21] mm^3^. The matrix sizes for all images include (128x, 256x, 320x, 512x). This work’s CNN models employ multimodal CT and MRI data. As mentioned previously, there are many forms of traditional MRI and CT scans that can form from different dyes, methods of taking the scan, or plane of section. Images were taken from axial, coronal, and sagittal sections. In addition, 20% of samples use C+ or Gadolinium contrast fluid.

### 4.2 Patch Sampling with Data Augmentation

As small datasets are prone to model overfitting and low accuracies, data augmentation was used in order to generate more data with the same images. The techniques used to create a variety of images from one sample include mirroring, random scaling, random rotations [-20,20] degrees, elastic deformations, resampling, random contrast, random brightness, and gamma correction. All data augmentation was accomplished alongside a python package from Medical Image Computing at the German Cancer Research Center (DKFZ) and the Applied Computer Vision Lab of the Helmholtz Imaging Platform^19^. All patches resulting from data processing are generated randomly during the training step of the model. Each batch has a foreground and a background class for easier image classification that are input to the training model. The foreground class allows for proper data augmentation through all transformations.

### 4.3 Image Preprocessing

The image preprocessing is independent of the CNN, and various steps were taken in order to optimize the input and resulting output for the training model. All preprocessing steps were done through the Advanced Normalization Tools (ANTs), Wellcome Centre for Human Neuroimaging (WCHN) CT normalization tools (CTseg), FMRIB’s Linear Image Registration Tool (FLIRT), and FMRIB’s Brain Extraction Tool (BET). Below is the list of methods employed to mitigate non-conformity:

- Image Registration: CT and MRI images are of different modalities and each have sub-weights. Image registration is vital in order to transform these different data sets into one dataset on one coordinate system.
- Bias field Correction: MRI images of all weights contain variance in the low frequency intensity in both the bias and gain fields. During this prepossessing step, bias was corrected in MRI images.
- Skull Stripping: Normal MRI and CT images contain not only brain matter, but the surrounding bone and tissue. Because the skull and other surrounding tissues introduce unfavorable bias and variables, skull stripping removes the skull from the end image while attempting to not distort brain matter.
- Image Normalization: Image intensity is standardized to a range of [0,1] and image size is normalized to (224, 224, 3).

### 4.4 Architecture Design and Attention Mechanisms

In this work, EfficientNetB0 was used as the backbone for CNN. This was used as the backbone for this work because of its employment of compound scaling instead of normal random scaling. Instead of the traditional method of balancing the scale in one dimension, compound scaling targets three different dimensions: width, depth, and image resolution. Under sufficient standardized testing by the authors of this CNN, it surpassed other methods without compound scaling in accuracy and efficiency. EfficientNetB0 was trained by over one million images and pre-trained weights from the ImageNet database, with 237 layers of varying filter sizes and over 11 million trainable parameters. EfficientNetB0 was used over its newer counterpart B7 because this lightwork CNN can still accomplish high accuracies.

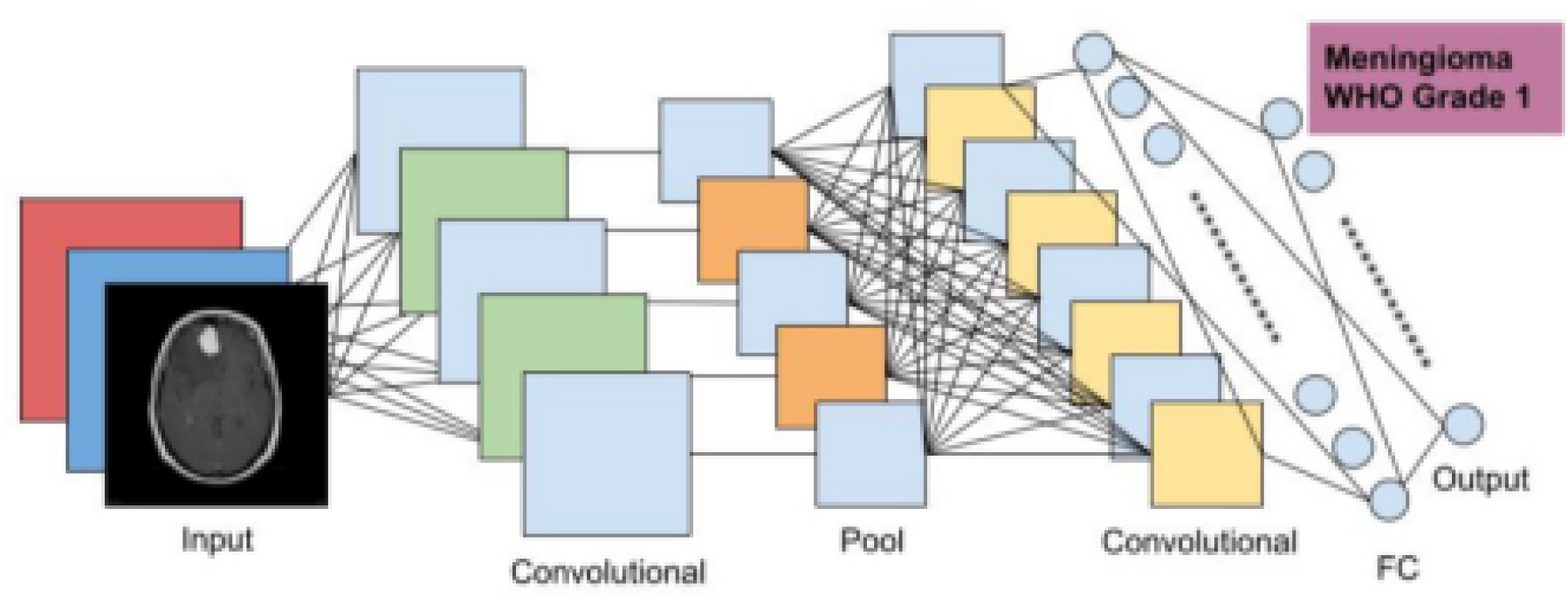

This model’s attention mechanisms are inherited and implemented from the keras.applications.EfficientNetB0, and therefore this work will not go in depth into encoder and decoder pathways nor linear transformations. EfficientNet uses composite coefficients to uniformly scale all dimensions of depth, width, and resolution. A simple self-attention network structure is trained in advance to process data images. This will enable the pre-training network to focus as much as possible on the data and image features with a large amount of information and a large gap before entering EfficientNet training. After that, the pre-training network will train the model.

The GlobalAveragePooling2D layer was included to reduce computational load during training by using average values instead of max values for pooling. Additionally, a dropout layer, which randomly omits some neurons at each step, helps prevent overfitting by making neurons more independent. Callbacks such as TensorBoard, ModelCheckpoint, and ReduceLROnPlateau were included in order to help monitor training, fix bugs, and prevent overfitting by implementing early stopping and adjusting the learning rate. Finally, a dense output layer classifies the image into either four or three classes, depending on the task, using the softmax function. Softmax is generally employed in multi-class situations, as it performs better than sigmoid, which is typically used in binary contexts.

### 4.5 Training Strategies

The CNN ran for 12 consecutive epochs, and ended there without any significant validation loss improvement. Two loss functions were used in these CNNs: class-average loss and Tversky Loss (FTL). Although typically deployed in segmentation contexts, FTL was adapted to fit classification outputs. Class-average loss was chosen in order to effectively demonstrate and improve overall CNN performance. The FTL was employed due to its strong capability in efficiently balancing false positive and false negative predictions, specifically with the inclusion of cysts as a minority in the dataset. FTL’s flexibility in the focal parameter also allows it to leverage original data image volumes to account for loss calculation. The values for FTL were: α = 0.7 and β = 0.3 for the Tversky index and γ = 2.0 as the focal parameter. These values attempt to limit the number of predicted false negatives by the model. In a medical context, false negatives (missing a tumor) are more serious than false positives, leading to a higher alpha value, which places weight on false negatives. The high value for the focal parameter ensures the model focuses on difficult outliers such as cyst growth or tumor volumes <3mL. During model training, the models were saved based on the loss function that resulted in the lowest validation loss.

All models were trained using two samples in a batch due to the large memory footprint. The models in this work use mini-batch sizes of 32 elements, with accumulated gradients to increase the size, which mimics the amount of gradients given larger batch sizes. Smaller batch sizes of up to 32 have demonstrated improvements in generalization and model quality^24^.

### 4.6 Data Analysis

In the study, a combination of statistical tests and qualitative analysis was employed to analyze the collected data. For the overall performance study, metrics such as f1, precision, recall, and accuracy were calculated and averaged across five folds for each tumor type and meningioma grade. These metrics were visualized using tables and graphs to summarize performance. Accuracy and loss analysis involved plotting validation and training accuracy and loss over 12 epochs, with error bars representing a 95% confidence interval, to statistically assess the model’s performance and consistency. Confusion matrices and ROC curves were generated using the SciKit Learn Python library to evaluate true positives, false positives, and AUC scores, providing a comprehensive statistical overview of the models’ classification capabilities. For qualitative analysis, attention maps were created using the GradCam TensorFlow extension, highlighting areas of high model attention on MRI images to ensure the model focused on relevant tumor regions. This combination of quantitative metrics and qualitative visualizations ensured a thorough evaluation of the model’s performance.

### 4.7 Ethical Considerations

Rights and consent were obtained for all imaging scans of patients and external programs used for data augmentation or alteration. Additionally, patient confidentiality was preserved; intimate patient information was a non-factor in data extraction.

## Data Availability

All data produced in the present study are available upon reasonable request to the authors

https://medpix.nlm.nih.gov/search?allen=true&allt=true&alli=true&query=meningioma

https://radiopaedia.org/search?lang=us&q=meningioma&scope=cases

https://github.com/sartajbhuvaji/brain-tumor-classification-dataset

## 5. ACKNOWLEDGEMENTS

This work was reviewed and researched alongside Harvard University PhD Doctor of Philosophy candidate Sharifa Sahai.

## Notes

### Competing Interest Statement

The authors have declared no competing interest.

### Funding Statement

This study did not receive any funding

### Author Declarations

The study used ONLY openly available human data that were originally located at: - https://radiopaedia.org/search?lang=us&q=meningioma&scope=cases - https://medpix.nlm.nih.gov/search?allen=true&allt=true&alli=true&query=meningioma - https://github.com/sartajbhuvaji/brain-tumor-classification-dataset

### Summary of Updates

All sections re-written to improve clarity and focus (references updated accordingly); references now follow American Association for the Advancement of Science (AAAS) guidelines, Tables 1 and 2 previously incorrectly contained data from different trials, and now contains data only within one trial. Images and sections reorganized; keywords section included in the abstract; ethical considerations sub-section included; figure captions updated for clarity.

